# Carriage of antimicrobial-resistant Enterobacterales among pregnant women and newborns in Amhara, Ethiopia

**DOI:** 10.1101/2023.05.29.23290490

**Authors:** Getnet Amsalu, Christine Tedijanto Wen, Olga Perovic, Addisalem Gebru, Bezawit M. Hunegnaw, Fisseha Tadesse, Marshagne Smith, Addisalem Fikre, Delayehu Bekele, Lisanu Taddesse, Grace Chan

## Abstract

**Background:** Infections are one of the most common causes of neonatal mortality, and maternal colonization has been associated with neonatal infection. Data on carriage of bacterial pathogens and mother-child transmission patterns in low- and middle-income countries is sparse.

**Methods:** We sought to quantify carriage prevalence of extended-spectrum-beta-lactamase (ESBL) - producing and carbapenem-resistant Enterobacterales (CRE) among pregnant women and their neonates and to characterize risk factors for carriage in a rural area of Amhara, Ethiopia. We sampled 211 pregnant women in their third trimester and/or labor/delivery and 159 of their neonates in the first week of life.

**Results:** We found that carriage of ESBL-producing organisms was fairly common (women: 22.3%, 95% CI: 16.8-28.5; neonates: 24.5%, 95% CI: 18.1-32.0), while carriage of CRE (women: 0.9%, 95% CI: 0.1-3.4; neonates: 2.5%, 95% CI: 0.7-6.3) was rare. Neonates whose mothers tested positive for ESBL-producing organisms were nearly twice as likely to also test positive for ESBL-producing organisms (38.7% vs. 21.1%, p-value: 0.06). Carriage of ESBL-producing organisms was also associated with woreda (district) of sample collection (Fisher exact test maternal p-value: <0.01; neonatal p-value: <0.01) and recent antibiotic use (maternal p-value: 0.55; neonatal p-value: 0.011).

**Conclusions:** Understanding carriage patterns of potential pathogens and antibiotic susceptibility among pregnant women and newborns in this region will help to inform local, data-driven recommendations to prevent and treat neonatal infections.

**Main point:** Carriage prevalence of ESBL-producing Enterobacterales was high among pregnant women and neonates in a rural area of Ethiopia, and neonates were more likely to test positive if their mother tested positive. This work informs prevention and treatment of neonatal infections.

## Introduction

Neonatal infections, particularly sepsis, meningitis, and pneumonia, are among the most common causes of mortality in the first 28 days of life, accounting for approximately 23% of 2.4 million neonatal deaths worldwide [1,2]. Treatment of infections is increasingly challenging in the context of antimicrobial resistance, with over 40% of neonatal sepsis cases estimated to have resistance or reduced susceptibility to both the first-(ampicillin/penicillin and gentamicin) and second-line (third-generation cephalosporins) treatments recommended by the World Health Organization (WHO) [3]. Better understanding of the pathways leading to neonatal infection could inform critical infection prevention strategies.

Maternal colonization has been shown to be associated with neonatal infection [4]. Shared pathogens between mothers and their newborns may be a result of vertical transmission at birth and/or common environmental exposures. This phenomenon has been well-studied in high-income settings, particularly for *Streptococcus agalactiae*/Group B *Streptococcus* (GBS), once a leading cause of neonatal infections in the United States. Resulting interventions such as prenatal screening and antibiotic prophylaxis during labor have decreased the incidence of invasive early-onset GBS disease by over 80% [5,6].

However, relatively little is known about maternal colonization and subsequent neonatal infections in low- and middle-income settings. Unfortunately, this often overlaps with settings with high burden of disease; for example, Sub-Saharan Africa has the highest levels of neonatal mortality at 27 deaths per 1000 live births (accounting for 43% of global newborn deaths) [2], and infections have been found to contribute a greater proportion of deaths in high neonatal mortality settings [7]. Data specific to these populations is greatly needed to inform relevant clinical guidelines.

Our study focused on pregnant women and newborns in the Birhan field site, a surveillance site focused on maternal and child health in North Shewa Zone, Amhara, Ethiopia [8]. In this cohort between November 2018 and December 2020, overall neonatal mortality was 3.1% of live births [9]. We estimated carriage prevalence of Gram-negative antibiotic-resistant organisms – extended-spectrum-beta-lactamase (ESBL) -producing organisms and carbapenem-resistant *Enterobacterales* (CRE). We also checked samples for the presence of GBS due to its global importance in neonatal infections. Using samples collected at antenatal care (ANC), labor/delivery, and during the first week of life, we assessed longitudinal carriage patterns during pregnancy and between mothers and their newborns. In addition, we studied the association between colonization with our organisms of interest and clinical and environmental risk factors.

Ethical clearance for this study was obtained from the Ethics Review Board (IRB) of Saint Paul’s Hospital Millennium Medical college, (Addis Ababa, Ethiopia) [PM23/274], Boston Children’s Hospital (Boston, United States) [IRB-P00028224], and Harvard T.H. Chan School of Public Health (Boston, United states) [IRB19-0991].

## Methods

### Study setting

We conducted a prospective cohort study at the Birhan field site, including 16 villages in Amhara Region, Ethiopia with a total population of 77,766. The catchment area is rural and semi-urban, covers both highland and lowland areas, and includes two different districts, Angolela Tera, and Kewot/Shewa Robit. The site includes a health and demographic surveillance system (HDSS), the Birhan HDSS, with house-to-house surveillance every three months to estimate morbidity and mortality outcomes among 17,108 women of reproductive age and 8,554 children under five years old. The site is a platform for community and facility-based research and training that was established in 2018 [8]. Nested in the site is an open pregnancy and birth cohort, the Birhan cohort, that enrolls approximately 2,000 pregnant women and their newborns per year with rigorous longitudinal follow-up over the first two years of life and household data linked with health facility information [10].

### Sample collection

Samples were collected from March to August 2022. Any pregnant woman who had an address in one of the Birhan catchment villages and visited any of the Birhan health facilities for an antenatal care (ANC) visit at >35 weeks or for labor/delivery was considered for enrollment in the study. Among women who were enrolled at ANC, follow-up samples at labor/delivery were collected at any of the Birhan health facilities.

Signed informed consent was obtained from all participants upon enrollment in the study. After receiving informed consent, the trained data collectors collected samples and administered the study questionnaire in the facility. Samples were excluded if complications such as premature rupture of membranes (PROM), antepartum hemorrhage, or genital ulcers were present. If a woman contributed her first samples at ANC, follow-up samples were requested at labor/delivery occurring in facilities. At each visit, Dacron swabs were used to collect two samples from each woman, one rectal and one vaginal. Neonatal samples were collected at day 6 after birth, coinciding with an existing follow-up visit in the ongoing Birhan MCH cohort. Either perirectal or stool samples were collected based on the family’s preference. Families could refuse to provide samples at any time.

Swabs were stored in facility refrigerators for up to 24 hours before being transported to Debre Birhan Hospital, the referral hospital for the North Shewa Zone. At Debre Birhan Hospital, samples were stored at 2-8°C for up to 3 weeks (typically <14 days) before shipment to the National Institute for Communicable Diseases (NICD) in South Africa. Standardized sample transportation techniques (e.g., triple packaging of samples) were used to maintain the viability of organisms and safety of the public and environment.

### Sample processing - antimicrobial susceptibility testing

Phenotypic identification and determination of antimicrobial susceptibility were performed at NICD, a division of National Health Laboratory Service (NHLS), South Africa. Interpretation of susceptibility breakpoints was according to Standard Clinical and Laboratory Standards Institute (CLSI) guidelines [11].

Samples received at NICD were processed upon receipt in the laboratory. A worksheet was created for each batch of processed swabs. Each swab was plated on following media: (1) MacConkey agar with an imipenem disc placed on the initial inoculum as an additional screen for carbapenem resistant organisms, (2) Colorex™ESBL for detection of Gram-negative bacteria producing Extended Spectrum Beta-Lactamase (ESBL), (3) Colorex™mSuperCARBA™ for detection and isolation of Carbapenem-Resistant *Enterobacterales* (CRE), and (4) Todd Hewitt enrichment broth for *Streptococcus agalactiae* (GBS). Plates and broths were incubated for 24-hours at 37°C.

Colorex™ agar plates were examined for colonies demonstrating various color changes according to the manufacturer’s instructions for each of the Colorex™ media used. MacConkey agar plates were examined for Gram negative bacteria growing close to the imipenem disc. Suspected colonies on Colorex™ media were plated out on MacConkey agar for identification and antimicrobial susceptibility testing. After 24-hours of incubation, colonies were identified using the MALDI TOF-MS Biotyper® system (Bruker Daltonics GmbH, Bremen, Germany) and AST performed on MicroScan® (Beckman Coulter, Inc., West Sacramento, CA, USA) using Microscan® Neg MIC Panel Type 44. The Todd Hewitt enrichment broth was sub-cultured on a blood agar plate and Colorex™StrepB agar plate and incubated for 24 hours at 37°C.

Suspected colonies from blood agar were identified on the MALDI Biotyper®. Suspected colonies from Colorex™StrepB agar were plated out on blood agar prior to identification and AST. Isolates of GBS underwent MIC testing using the Sensititre (ThermoFisher). *Streptococcus* species STP6F Trek panel as well as Kirby Bauer disc susceptibility testing. Isolates of ESBL-producing organisms, CRE and GBS, were stored in tryptic soy broth with 10% glycerol in -70°C.

### Sample processing - molecular testing

Molecular testing was also conducted to verify the presence of ESBL genes among isolates identified as ESBL-positive by antimicrobial susceptibility testing profiles to the third and fourth generation of cephalosporins identified by MicroScan®. The DNA was extracted using a crude boiling method at 95°C for 25 minutes. The DNA templates were then tested for the presence of extended-spectrum beta-lactamase (ESBL) genes such as *bla*_*-*TEM_, *bla*_*-*SHV_ and *bla*_*-*CTXM_ using the LightCycler 480 instrument (Roche Applied Science, Germany) and LightCycler 480 Probes Master kit (Roche Diagnostics, USA) in a real-time polymerase chain reaction (PCR) assay. The *bla*_*-*TEM_, *bla*_*-*SHV_ and *bla*_*-*CTXM_ genes were amplified by multiplex real-time PCR using the primers and probes shown in **Supplementary Table 1**. For *bla*_*-*TEM and_ *bla*_*-*SHV,_ the reaction conditions were 10μM primers, 2μM probes, denaturation for 95°C for 5 minutes, and then 45 cycles of 95°C for 10 seconds, 55°C for 30 seconds and 72°C for 1 second. The *bla*_*-*CTXM_ PCR was a multiplex assay targeting *bla*_*-*CTXM_ group M1 and M2-9 using primer and probe sequences as described previously [12].

### Sample processing - whole genome sequencing

Whole genome sequencing was conducted to verify the presence of CRE genes among isolates identified as CRE-positive by antimicrobial susceptibility testing profiles to carbapenems identified by MicroScan®. Genomic DNA (gDNA) was extracted using QIAamp DNA mini kit (Qiagen, TX, USA) following the manufacturer’s instructions. The concentrations of the extracted gDNA were determined using Qubit 4.0 fluorometer (ThermoFisher Scientific, Waltham, MA, USA). Multiplexed paired-end libraries were prepared using the Illumina DNA Prep kit (Illumina, San Diego, CA, USA). Sequencing was performed on Illumina NextSeq 550 platform (Illumina, San Diego, CA, USA) (2x 150bp) with 100x coverage at the NICD Sequencing Core Facility, NHLS, South Africa.

Raw paired-end reads were analysed using the Jekesa pipeline (v1.0; https://github.com/stanikae/jekesa). Briefly, Trim Galore! (v0.6.2; https://github.com/FelixKrueger/TrimGalore) was used to filter the generated sequence raw reads (Q >30 and length >50 bp). De novo assembly and optimization of the contigs were performed using SPAdes v3.13 and Shovill (v1.1.0; https://github.com/tseemann/shovill), respectively. The multilocus sequence typing (MLST) profiles were determined using the MLST tool (version 2.16.4; https://github.com/tseemann/mlst). Assembly metrics were calculated using QUAST (v5.0.2; http://quast.sourceforge.net/quast).

Whole-genome single nucleotide polymorphism (SNP) differences were determined with a reference-free approach using the SKA toolkit [13]. Antibiotic resistance profiles and virulence genes were predicted using ABRicate (version 1.0.1; https://github.com/tseemann/ABRicate), against the Comprehensive Antibiotic Resistance Database (CARD), ResFinder - Center for Genomic Epidemiology (CGE) database [14], NCBI AMRFinderPlus [15], and Virulence Factor Database (VFDB) [16] implemented in the Jekesa pipeline. Pathogen Watch (https://pathogen.watch/) was used to construct the phylogenetic tree [Newick (NWK) file]. The exported NWK file was used in Microreact (https://microreact.org/showcase) to visualize and edit the phylogenetic tree. The assembled genome files were submitted to the National Center for Biotechnology Information GenBank and are available under BioProject number: PRJNA819852[RM1].

### Statistical analysis

Carriage prevalence at each time point was calculated as the proportion of positive samples over the number of total samples. We also evaluated the proportion of women and children who tested positive for each organism in any sample and at any time point. Due to relatively small sample sizes, the Fisher exact test was used to statistically evaluate associations between carriage outcomes and potential risk factors, and an alpha level of 0.05 was used to determine significance. Data cleaning and analysis were conducted in R (version 4.2.0) [17].

## Results

In total, 460 samples were collected from 211 women at ANC and/or delivery, and 159 samples were collected from neonates at day 6 after birth (**Figure 1**). Of the 171 women who gave their first sample at an antenatal care appointment, 19 of them also gave a second sample at delivery and 120 of their children provided samples at Day 6 (123 total neonates due to 3 pairs of twins). Many women were not followed up at labor/delivery due to delivering at home (26/152, 17.1%) or at night (106/152, 69.7%) when study data collectors were not present, or other unknown reasons (20/152, 13.2%). Of the 40 women who gave their first sample at delivery, 35 of their children provided samples at Day 6 (36 total neonates due to 1 pair of twins). Overall, 56 women who gave samples at ANC and/or labor/delivery did not have neonates who contributed to the study due to ending of the study period (26/56, 46.4%), unavailability of data collector (11/56, 19.6%), missed visits due to social unrest (5/56, 8.9%), family refusal (4/56, 7.1%), migration out of the study area (4/56, 7.1%), stillbirth or early neonatal death (4/56, 7.1%), or sample rejection at the lab (2/56, 3.6%).

**Figure 1.**
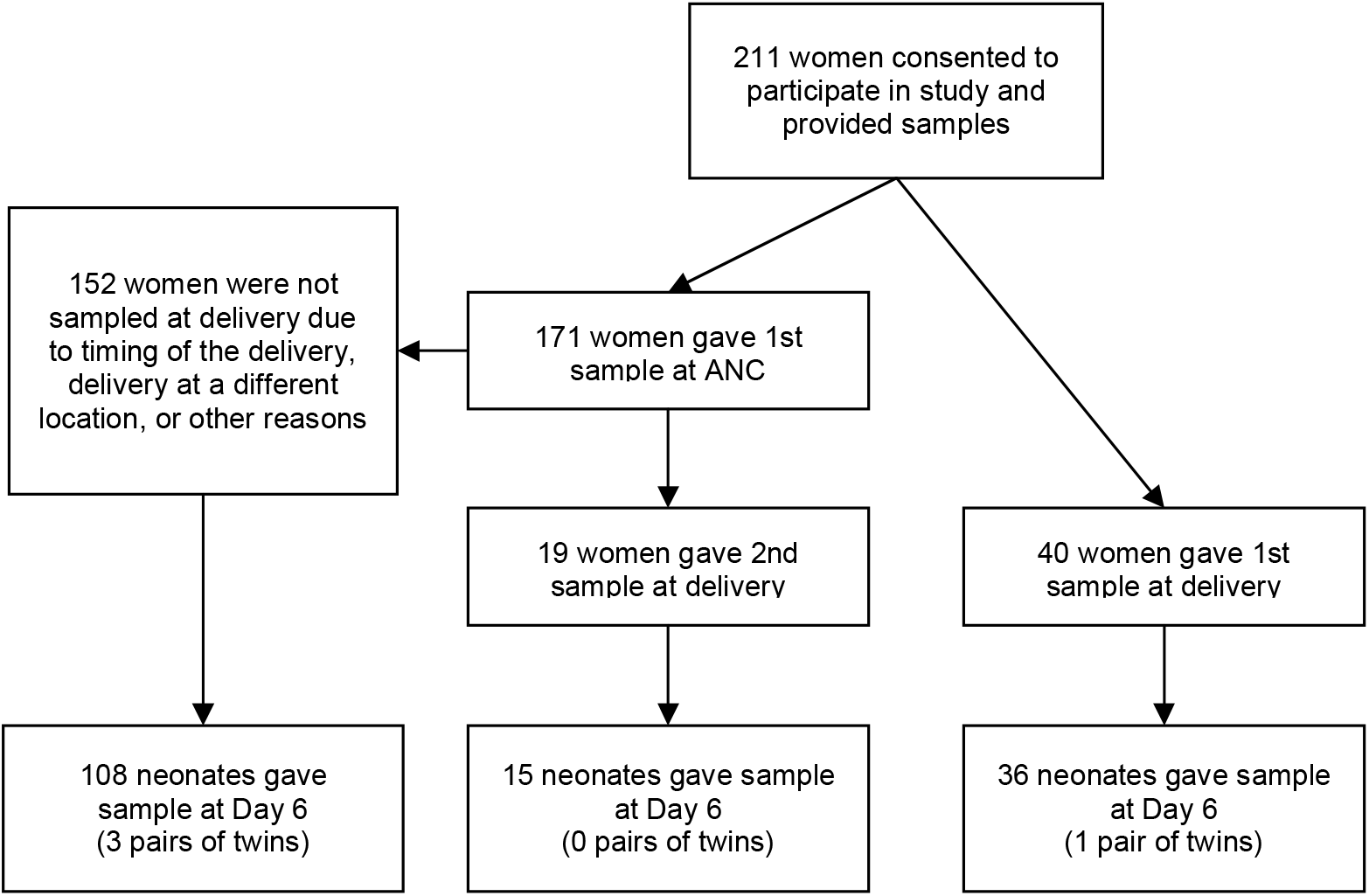
Study flow chart.

**Figure 2.**
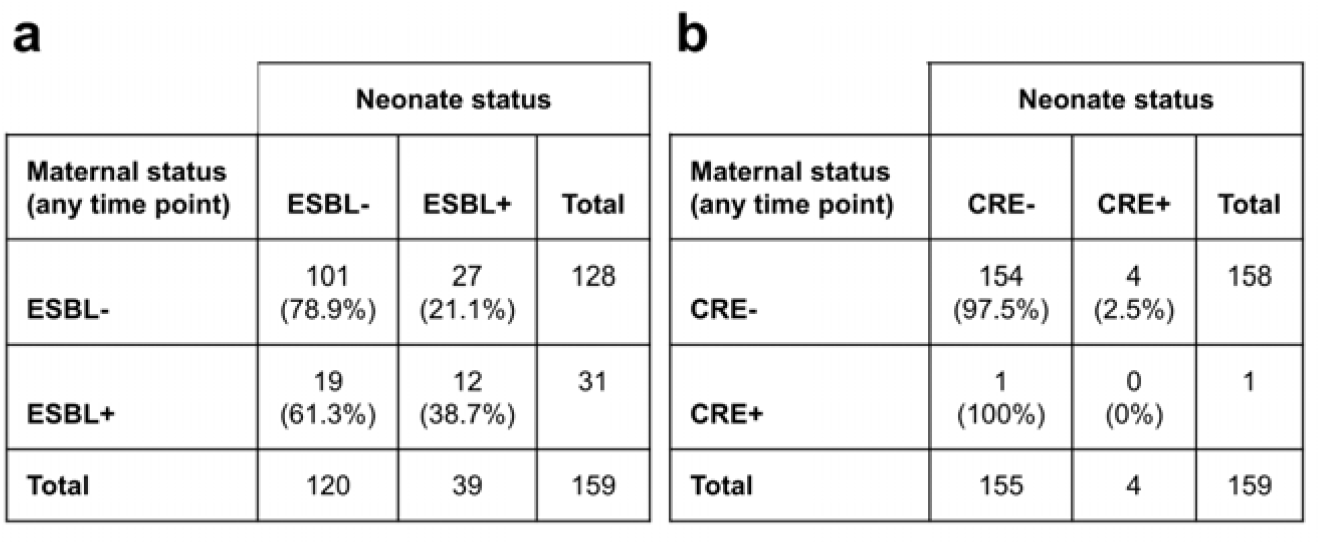
2×2 tables comparing carriage of ESBL-producing organisms (a) and CRE (b) in mothers at any point during pregnancy and their neonates.

Overall, 22.3% of women were positive for ESBL-producing organisms considering any time point or sample type and 0.9% were positive for CRE (**Table 1**). ESBL was more common in rectal swabs compared to vaginal swabs (19.9% vs. 0.6% at ANC and 22.0% vs. 0% at labor/delivery). Carriage prevalence among neonates followed a similar overall pattern, with CRE being much more rare than ESBL. Across all sample types, 24.5% of neonates were positive for ESBL-producing organisms, and 2.5% were positive for CRE. The majority of ESBL-producing isolates were *Escherichia coli* (82/102, 80.3%), followed by *Klebsiella pneumoniae* (13/102, 12.7%) and *Enterobacter cloacae* (7/102, 6.9%). In most cases, one ESBL-producing organism was isolated per specimen; two different organisms were isolated from the same swab in 15 out of 87 cases (17.2%). The majority of ESBL-producing isolates carried the CTX-M1 gene only (30.7%) or TEM and CTX-M-1 (42.6%) (**Supplementary Table 2**).

**Table 1.** Carriage prevalence of ESBL-producing organisms and CRE among mothers and neonates by time point and sampling site.

| Sample type | Time point | Sample size | ESBL % (95% CI) | CRE % (95% CI) |
| --- | --- | --- | --- | --- |
| **Maternal samples** |  |  |  |  |
| Unique women, any sample type | Any | 211 | **22.3** (16.8, 28.5) | **0.9** (0.1, 3.4) |
| Rectal | ANC | 171 | **19.9** (14.2, 26.7) | **0** (0, 2.1) |
| Vaginal | ANC | 171 | **0.6** (0.01, 3.2) | **0** (0, 2.1) |
| Rectal | Labor/delivery | 59 | **22.0** (12.3, 34.7) | **3.4** (0.4, 11.7) |
| Vaginal | Labor/delivery | 59 | **0** (0, 6.1) | **0** (0, 6.1) |
| **Neonatal samples** |  |  |  |  |
| Unique children, any sample type | Day 6 | 159 | **24.5** (18.1, 32.0) | **2.5** (0.7, 6.3) |
| Perirectal | Day 6 | 73 | **24.7** (15.3, 36.1) | **2.7** (0.3, 9.5) |
| Stool | Day 6 | 86 | **24.4** (15.8, 34.9) | **2.3** (0.3, 8.1) |

**Table 2.** Associations between sample, clinical, and environmental characteristics and carriage outcomes in pregnant women. Unless otherwise noted, the unit of analysis was the individual-visit (n = 171 ANC visits + 59 labor/delivery visits = 230); the unit was considered positive if either the rectal or vaginal swab returned a positive result.

| Variable | Sample size | ESBL |  |  | CRE |  |  |
| --- | --- | --- | --- | --- | --- | --- | --- |
|  |  | Positive n (%) | Negative n (%) | p-value | Positive n (%) | Negative n (%) | p-value |
| Sample characteristics |  |  |  |  |  |  |  |
| Sample type<sup>a</sup> |  |  |  |  |  |  |  |
| Rectal | 230 | 47 (20.4) | 183 (79.6) | <0.01 | 2 (0.9) | 228 (99.1) | 0.5 |
| Vaginal | 230 | 1 (0.4) | 229 (99.6) |  | 0 (0) | 230 (100) |  |
| Woreda of sample collection |  |  |  |  |  |  |  |
| Angolela / Tera | 142 | 11 (7.7) | 131 (92.3) | <0.01 | 0 (0) | 142 (100) | 0.15 |
| Kewot | 88 | 37 (42.0) | 51 (58.0) |  | 2 (2.3) | 86 (97.7) |  |
| Facility type of sample collection |  |  |  |  |  |  |  |
| Health center | 196 | 35 (17.9) | 161 (82.1) | 0.011 | 2 (1.0) | 194 (99.0) | 1 |
| Hospital | 34 | 13 (38.2) | 21 (61.8) |  | 0 (0) | 34 (100) |  |
| Clinical characteristics |  |  |  |  |  |  |  |
| Sample collected before membrane rupture (labor/delivery samples only) |  |  |  |  |  |  |  |
| Yes | 43 | 9 (20.9) | 34 (79.1) | 0.83 | 0 (0) | 43 (100) | 0.12 |
| No | 14 | 4 (28.6) | 10 (71.4) |  | 2 (14.3) | 12 (85.7) |  |
| Missing | 2 | 0 (0) | 2 (100) |  | 0 (0) | 2 (100) |  |
| Received antibiotics within 3 months before sample collection |  |  |  |  |  |  |  |
| Yes | 22 | 6 (27.2) | 16 (72.8) | 0.55 | 1 (4.5) | 21 (95.5) | 0.21 |
| No | 205 | 42 (20.5) | 163 (79.5) |  | 1 (0.5) | 204 (99.5) |  |
| Missing | 3 | 0 (0) | 3 (100) |  | 0 (0) | 3 (100) |  |
| Hospitalized within 3 months before sample collection |  |  |  |  |  |  |  |
| Yes | 2 | 1 (50.0) | 1 (50.0) | 0.45 | 0 (0) | 2 (100) | 1 |
| No | 225 | 47 (20.9) | 178 (79.1) |  | 2 (0.9) | 223 (99.1) |  |
| Missing | 9 | 0 (0) | 3 (100) |  | 0 (0) | 3 (100) |  |
| Environmental factors |  |  |  |  |  |  |  |
| Residence type |  |  |  |  |  |  |  |
| Rural | 173 | 38 (22.0) | 135 (78.0) | 0.57 | 2 (1.2) | 171 (98.8) | 1 |
| Urban | 57 | 10 (17.5) | 47 (82.5) |  | 0 (0) | 57 (100) |  |
| Household size |  |  |  |  |  |  |  |
| 4 individuals (median) or fewer | 112 | 30 (26.8) | 82 (73.2) | 0.11 | 1 (0.9) | 111 (99.1) | 1 |
| More than 4 individuals | 69 | 10 (14.5) | 59 (85.5) |  | 1 (1.4) | 68 (98.6) |  |
| Missing | 49 | 8 (16.3) | 41 (83.7) |  | 0 (0) | 49 (100) |  |
| Type of toilet at home |  |  |  |  |  |  |  |
| Pit latrine with slab | 32 | 3 (9.4) | 29 (90.6) | 0.14 | 0 (0) | 32 (100) | 1 |
| Pit latrine without slab | 74 | 21 (28.4) | 53 (71.6) |  | 1 (1.4) | 73 (98.6) |  |
| Other | 75 | 16 (21.3) | 59 (78.7) |  | 1 (1.3) | 74 (98.7) |  |
| Missing | 49 | 8 (16.3) | 41 (83.7) |  | 0 (0) | 49 (100) |  |
| Drinking water source at home |  |  |  |  |  |  |  |
| Piped to home or nearby | 50 | 11 (22.0) | 39 (78.0) | 0.44 | 1 (2.0) | 49 (98.0) | 0.71 |
| Public tap | 58 | 16 (27.6) | 42 (72.4) |  | 0 (0) | 58 (100) |  |
| Well, spring, or surface water | 71 | 13 (18.3) | 58 (81.7) |  | 1 (1.4) | 70 (98.6) |  |
| Missing | 51 | 8 (15.7) | 43 (84.3) |  | 0 (0) | 51 (100) |  |
| Flooring material at home |  |  |  |  |  |  |  |
| Natural | 121 | 20 (16.5) | 101 (83.5) | 0.027 | 2 (1.7) | 119 (98.3) | 1 |
| Finished | 60 | 20 (33.3) | 40 (66.7) |  | 0 (0) | 60 (100) |  |
| Missing | 49 | 8 (16.3) | 41 (83.7) |  | 0 (0) | 49 (100) |  |
| Own livestock |  |  |  |  |  |  |  |
| Yes | 89 | 17 (19.1) | 72 (80.9) | 0.44 | 1 (1.1) | 88 (98.9) | 1 |
| No | 92 | 23 (25.0) | 69 (75.0) |  | 1 (1.1) | 91 (98.9) |  |
| Missing | 49 | 8 (16.3) | 41 (83.7) |  | 0 (0) | 49 (100) |  |
| Domestic animals cohabitate with humans |  |  |  |  |  |  |  |
| Yes | 42 | 3 (7.1) | 39 (92.9) | 0.022 | 1 (2.4) | 41 (97.6) | 0.23 |
| No | 120 | 26 (21.7) | 94 (78.3) |  | 0 (0) | 120 (100) |  |
| Missing | 68 | 19 (27.9) | 49 (72.1) |  | 1 (1.5) | 67 (98.5) |  |

<sup>a</sup>The unit of analysis for this variable was each sample.

**Table 3.** Associations between sample, clinical, and environmental characteristics and carriage outcomes in neonates.

| Outcomes in neonates: |  |  |  |  |  |  |  |
| --- | --- | --- | --- | --- | --- | --- | --- |
| Variable | Sample size | ESBL |  |  | CRE |  |  |
|  |  | Positive n (%) | Negative n (%) | p-value | Positive n (%) | Negative n (%) | p-value |
| Sample characteristics |  |  |  |  |  |  |  |
| Sample type |  |  |  |  |  |  |  |
| Perirectal | 73 | 18 (24.7) | 55 (75.3) | 1 | 2 (2.7) | 71 (97.3) | 1 |
| Stool | 86 | 21 (24.4) | 65 (75.6) |  | 2 (2.3) | 84 (97.7) |  |
| Woreda of sample collection |  |  |  |  |  |  |  |
| Angolela / Tera | 101 | 12 (11.9) | 89 (88.1) | <0.01 | 2 (2.0) | 99 (98.0) | 0.62 |
| Kewot | 58 | 27 (46.6) | 31 (53.4) |  | 2 (3.4) | 56 (96.6) |  |
| Location of sample collection |  |  |  |  |  |  |  |
| Community | 141 | 32 (22.7) | 109 (77.3) | 0.16 | 1 (0.7) | 140 (99.3) | <0.01 |
| Health center or hospital | 1 | 1 (100) | 0 (0) |  | 1 (100) | 0 (0) |  |
| Other | 1 | 0 (0) | 1 (100) |  | 0 (0) | 1 (100) |  |
| Missing | 16 | 6 (37.5) | 10 (62.5) |  | 2 (12.5) | 14 (87.5) |  |
| Clinical characteristics |  |  |  |  |  |  |  |
| Preterm birth |  |  |  |  |  |  |  |
| Yes | 19 | 4 (21.1) | 15 (78.9) | 0.70 | 1 (5.3) | 18 (94.7) | 0.48 |
| No | 135 | 33 (24.4) | 102 (75.6) |  | 3 (2.2) | 132 (97.8) |  |
| Missing | 5 | 2 (40.0) | 3 (60.0) |  | 0 (0) | 5 (100) |  |
| Location of birth |  |  |  |  |  |  |  |
| Community | 22 | 3 (13.6) | 19 (86.4) | <0.01 | 0 (0) | 22 (100) | 0.09 |
| Health center | 98 | 17 (17.3) | 81 (82.7) |  | 1 (1.0) | 97 (99.0) |  |
| Hospital | 33 | 17 (51.5) | 16 (48.5) |  | 3 (9.1) | 30 (90.9) |  |
| Missing | 6 | 2 (33.3) | 4 (66.7) |  | 0 (0) | 6 (100) |  |
| Mother received antibiotics at labor/delivery |  |  |  |  |  |  |  |
| Yes | 24 | 11 (45.8) | 13 (54.2) | 0.036 | 2 (8.3) | 22 (91.7) | 0.15 |
| No | 110 | 22 (20.0) | 88 (80.0) |  | 2 (1.8) | 108 (98.2) |  |
| Missing | 25 | 6 (24.0) | 19 (76.0) |  | 0 (0) | 25 (100) |  |
| Received antibiotics after birth |  |  |  |  |  |  |  |
| Yes | 9 | 6 (66.7) | 3 (33.3) | 0.011 | 2 (22.2) | 7 (77.8) | 0.03 |
| No | 141 | 32 (22.7) | 109 (77.3) |  | 2 (1.4) | 139 (98.6) |  |
| Missing | 9 | 1 (11.1) | 8 (88.9) |  | 0 (0) | 9 (100) |  |
| Received care in facility after birth |  |  |  |  |  |  |  |
| Yes | 2 | 2 (100) | 0 (0) | 0.046 | 0 (0) | 2 (100) | 0.12 |
| No | 140 | 31 (22.1) | 109 (77.9) |  | 2 (1.4) | 138 (98.6) |  |
| Unknown | 1 | 0 (0) | 1 (100) |  | 0 (0) | 1 (100) |  |
| Missing | 16 | 6 (37.5) | 10 (62.5) |  | 2 (12.5) | 14 (87.5) |  |
| **Environmental factors** |  |  |  |  |  |  |  |
| ***Residence type*** |  |  |  |  |  |  |  |
| Rural | 128 | 34 (26.6) | 94 (73.4) |  | 3 (2.3) | 125 (97.7) |  |
| Urban | 31 | 5 (16.1) | 26 (83.9) | 0.26 | 1 (3.2) | 30 (96.8) | 0.58 |
| ***Household size*** |  |  |  |  |  |  |  |
| 4 individuals or fewer | 72 | 20 (27.8) | 52 (72.2) |  | 4 (5.6) | 68 (94.4) |  |
| More than 4 individuals | 55 | 11 (20.0) | 44 (80.0) |  | 0 (0) | 55 (100) |  |
| Missing | 32 | 8 (25.0) | 24 (75.0) | 0.62 | 0 (0) | 32 (100) | 0.14 |
| ***Type of toilet at home*** |  |  |  |  |  |  |  |
| Pit latrine with slab | 22 | 6 (27.3) | 16 (72.7) |  | 1 (4.5) | 21 (95.5) |  |
| Pit latrine without slab | 52 | 12 (23.1) | 40 (76.9) |  | 0 (0) | 52 (100) |  |
| Other | 53 | 13 (24.5) | 40 (75.5) |  | 3 (5.7) | 50 (94.3) |  |
| Missing | 32 | 8 (25.0) | 24 (75.0) | 0.99 | 0 (0) | 32 (100) | 0.15 |
| ***Drinking water source at home*** |  |  |  |  |  |  |  |
| Piped to home or nearby | 25 | 5 (20.0) | 20 (80.0) |  | 0 (0) | 25 (100) |  |
| Public tap | 47 | 14 (29.8) | 33 (70.2) |  | 1 (2.1) | 46 (97.9) |  |
| Well, spring, or surface water | 53 | 11 (20.8) | 42 (79.2) |  | 3 (5.7) | 50 (94.3) |  |
| Missing | 34 | 9 (26.5) | 25 (73.5) | 0.70 | 0 (0) | 34 (100) | 0.48 |
| ***Flooring material at home*** |  |  |  |  |  |  |  |
| Natural | 88 | 21 (23.9) | 67 (76.1) |  | 3 (3.4) | 85 (96.6) |  |
| Finished | 39 | 10 (25.6) | 29 (74.4) |  | 1 (2.6) | 38 (97.4) |  |
| Missing | 32 | 8 (25.0) | 24 (75.0) | 0.97 | 0 (0) | 32 (100) | 0.81 |
| ***Own livestock*** |  |  |  |  |  |  |  |
| Yes | 64 | 12 (18.8) | 52 (81.2) |  | 1 (1.6) | 63 (98.4) |  |
| No | 63 | 19 (30.2) | 44 (69.8) |  | 3 (4.8) | 60 (95.2) |  |
| Missing | 32 | 8 (25.0) | 24 (75.0) | 0.31 | 0 (0) | 32 (100) | 0.43 |
| ***Domestic animals cohabitate with humans*** |  |  |  |  |  |  |  |
| Yes | 25 | 5 (20.0) | 20 (80.0) |  | 0 (0) | 25 (100) |  |
| No | 94 | 22 (23.4) | 72 (76.6) | 0.66 | 2 (2.1) | 92 (97.9) | 0.62 |
| Missing | 40 | 12 (30.0) | 28 (70.0) |  | 2 (5.0) | 38 (95.0) |  |

All 16 CRE-positive isolates harbored more than one resistance gene (**Supplementary Table 3**). Among these sequenced isolates, 11 were identified as *E. coli*, 3 as *K. pneumoniae*, and 2 as *E. cloacae*. The majority of isolates (94%) carried at least one beta-lactamase gene, in different combinations; the most common was CTX-M-15 (68.8%), followed by TEM-1 (43.8%). Five isolates harbored the carbapenemase OXA-1 gene (31.3%), and one had a point mutation in the ompK37 locus (6.3%), which can confer resistance to carbapenems and other beta-lactams through reduced permeability.

There was an association between maternal and neonatal carriage of ESBL-producing organisms, with 21.1% of women who were never ESBL-positive having children who were ESBL-positive in the first week of life compared to 38.7% of ESBL-positive women (Fisher exact test p-value = 0.06). Among the twelve mother-baby pairs that were ESBL-positive, all of the women carried *E. coli*; 8 of the neonates also had only *E. coli*, 1 neonate had *E. coli* and *E. cloacae*, 1 neonate had *E. coli* and *K. pneumoniae*, 1 neonate had only *E. cloacae*, and 1 neonate had only *K. pneumoniae*. Only one woman whose neonate was also sampled tested positive for CRE. In addition, 4 neonates tested positive for CRE despite having mothers who were not positive during pregnancy. Results were similar when focusing only on maternal samples at labor/delivery (n = 51 mother-baby pairs; ESBL p-value <0.01) (**Supplementary Figure 1**). Among the 4 women whose neonates did not contribute samples due to stillbirth or early neonatal death, 2 were positive for ESBL and 1 was positive for CRE at ANC or labor/delivery.

Maternal carriage of ESBL-producing organisms was significantly associated with sample type (20.4% rectal vs. 0.4% vaginal), woreda of sample collection (42.0% in Kewot vs. 7.7% in Angolela/Tera), and location of sample collection (38.2% in hospitals vs. 17.9% in health centers). CRE was also only found on rectal swabs. Although none of the clinical characteristics reached statistical significance, there is evidence that recent antibiotic use is associated with carriage of ESBL-producing organisms (27.2% among women who took antibiotics in the last 3 months vs. 20.5% among women who did not) and CRE (4.5% vs. 0.5%). The most common antibiotics taken were amoxicillin (9/22, 40.9%), cephalexin (3/22, 13.6%), and chloramphenicol (2/22, 9.1%). Maternal carriage of ESBL-producing organisms was positively associated with finished household floors and lack of animal cohabitation. Other environmental characteristics such as household size, toilet type, water source, and livestock ownership were not associated with carriage of ESBL-producing organisms.

As with the maternal samples, ESBL-producing organisms were more common among neonates in Kewot (46.6%) compared to Angolela/Tera (11.9%). In addition, early exposure to antibiotics, either from maternal exposure at labor/delivery or after birth, was associated with neonatal carriage of ESBL-producing organisms and CRE. Among the 9 neonates who received antibiotics in the first week of life, 66.7% and 22.2% tested positive for ESBL-producing organisms and CRE, respectively, compared to 22.7% and 1.4% among neonates who did not receive antibiotics; in most cases (7/9, 77.8%), ampicillin was administered in combination with at least one other antibiotic, typically gentamicin (5/7, 71.4%), but ceftriaxone, cefotaxime, ceftazidime, vancomycin, metronidazole, and cloxacillin were also administered to at least one neonate each. A much higher proportion of neonates born in the hospital tested positive for ESBL-producing organisms compared to neonates born in the community (51.5% vs. 13.6%). In addition, all 4 neonates who tested positive for CRE were born in a health center or hospital. Both neonates who received care in a health facility in the first week of life also tested positive for ESBL-producing organisms. None of the environmental characteristics we explored were associated with neonatal carriage outcomes.

One case of neonatal sepsis occurred among the infants in our study. This case occurred prior to the Day 6 visit, which took place in the hospital, and was treated with the standard regimen of ampicillin and gentamicin. This neonate tested positive for both CRE and ESBL at Day 6, but we cannot determine whether these organisms were involved in the infection, arose during treatment, or were acquired later.

### Carriage prevalence of Group B Streptococcus

All samples were also assessed for presence of GBS. Overall, we found that carriage prevalence of GBS was rare in our study population. Only one woman tested positive for GBS at any time point (0.5%, at ANC). Two neonates tested positive (1.3%), though neither of their mothers tested positive. Both positive neonates were born in a health facility.

## Discussion

Among 211 pregnant women and 159 of their neonates in Amhara, Ethiopia, carriage prevalence of ESBL-producing organisms was around 25%, and carriage prevalence of CRE and GBS were less than 3%. Carriage of ESBL-producing organisms was associated with recent exposure to antibiotics and healthcare settings, though these relationships were not statistically significant for the maternal samples. In addition, ESBL-producing organisms were more common in rectal compared to vaginal maternal samples and in Kewot, where the climate is warmer and rainier compared to Angolela/Tera. Maternal carriage of ESBL-producing organisms at ANC or labor/delivery was associated with neonatal carriage in the first week of life.

There were 27, 4, and 2 newborns who tested positive for ESBL-producing organisms, CRE, and GBS, respectively, but had mothers who did not test positive during pregnancy. In most cases (21 ESBL, 2 CRE, 1 GBS), the women contributed samples at ANC only; it is possible that they acquired the organisms later in pregnancy and would have tested positive at labor/delivery. However, the neonates may also have become colonized through separate pathways. Investigation of such pathways may be important to inform future interventions to prevent neonatal carriage and potential infection with these pathogens. Many of the neonates with either ESBL-producing organisms (10/27, 37%) or CRE (3/4, 75%) were exposed to antibiotics during labor/delivery or after birth, which may have selected for antibiotic-resistant organisms. Many of the neonates were also born in a hospital (11/27 or 41% for ESBL, 3/4 or 75% for CRE) or health center (11/27 or 41% for ESBL, 1/4 or 25% for CRE), where they may have been exposed to antibiotic-resistant organisms.

To our knowledge, this was the first study to measure asymptomatic carriage prevalence of antimicrobial-resistant organisms and GBS in rural Ethiopia. The colonization prevalence estimates in our study were lower than existing reports from Ethiopia, a difference that may be partially attributable to study population. For example, a meta-analysis found that the prevalence of ESBL-producing Gram-negative bacteria across 17 studies in Ethiopia was 48.9% (95% CI: 40.2, 57.75) [19] compared to 20-25% in our study. Nearly all studies in the meta-analysis used clinical samples from patients visiting or staying in a health facility, which may have led to higher prevalence estimates. CRE has not been frequently measured in Ethiopia, but two facility-based studies have reported prevalence estimates of 2.4% (NICU) and 12.12% (inpatients and outpatients <15 years old) for carbapenemase-producing Gram-negative bacteria in clinical samples from children [20,21], compared to our estimate of 2.5% among children in the community. Finally, a recent systematic review of 16 studies in Ethiopia estimated that GBS prevalence in pregnant women was 16% (95% CI: 13, 20) [18] compared to 0.5% in our study. However, the majority of studies were conducted in large cities, where prevalence may differ from rural regions.

Existing studies of colonization in pregnant women and their babies have focused on GBS; a meta-analysis of 31 studies of mothers colonized with GBS estimated that 38.9% (95% CI: 29.6, 48.2) of their newborns had surface GBS colonization [22], similar to our estimate of 38.7% for ESBL-producing organisms shared between mothers and children. We also explored clinical and environmental risk factors that were previously shown to be associated with AMR colonization [23], but likely did not have enough power to detect significant associations in some cases. As expected, prior antibiotic use emerged as a risk factor for carriage of ESBL-producing organisms. This association may not have been significant for pregnant women as they could have taken antibiotics up to 3 months prior to sampling, while for neonates, antibiotic exposure occurred within a week prior to sampling. ESBL-producing organisms were more common among both pregnant women and neonates in Kewot woreda, which is warmer, rainier, and at lower altitude than Angolela/Tera. This finding may also reflect differences in healthcare seeking behavior, antibiotic use, and/or infection prevalence between the woredas, though further research is needed to measure these factors. Maternal carriage of ESBL-producing organisms was also associated with household flooring material and cohabitation with domestic animals.

Prevalence was higher among women with finished floors compared to natural floors and among households in which animals did not share rooms with humans. These observations run counter to our expectations and may be the result of confounding; for example, women with higher socioeconomic status may be more likely to have finished floors and also more likely to receive care at hospitals and be exposed to antibiotic-resistant organisms.

Our study has many strengths. We leveraged the well-characterized maternal and child health cohort of the Birhan HDSS in Amhara, Ethiopia. Due to the existing infrastructure, we were able to follow mother-baby pairs longitudinally from ANC to after birth. As data is available on all households and pregnant women and children are followed up frequently as part of the cohort, we had detailed information on clinical, environmental, socioeconomic and demographic factors. Sample processing and validation were conducted by a trained team at NICD in South Africa. PCR and WGS techniques were used to identify antibiotic resistance genes; this type of data is sparse for rural, LMIC settings.

However, our study was also subject to several limitations. First, we were unable to collect samples for many women at labor/delivery due to time of delivery (e.g., delivering at night when our data collectors were not available to collect samples), or location of delivery. Although carriage at labor/delivery would likely be the most indicative measure of neonatal carriage, the samples collected at late-term antenatal care (after 35 weeks) reflect the typical time period for screening of GBS [24]. Next, due to the small sample size, we were unable to control for confounding through multivariate analyses; however, the associations reported here may contribute to hypotheses for future research. Lastly, we limited our study to neonatal colonization rather than neonatal infection, as assessing this relatively rare outcome prospectively would have required a very large sample size. However, because we are nested within an ongoing surveillance cohort, we were able to observe one case of neonatal sepsis that occurred in our study population.

## Conclusions

In a rural area of Amhara, Ethiopia, maternal and neonatal carriage of ESBL-producing organisms was around 25%, and carriage of CRE and GBS were very rare. Neonates whose mothers tested positive for ESBL-producing organisms at late-term antenatal care or labor/delivery were roughly twice as likely to test positive in the first week after birth. Based on our findings, future carriage studies of CRE and ESBL-producing organisms can focus on rectal swabs, the source of nearly all isolates, over vaginal swabs. In some cases, neonates carried ESBL-producing organisms, CRE, or GBS even though their mothers did not, potentially due to antibiotic use or exposure in health facilities. Increased monitoring of ESBL-producing organisms may be warranted, particularly in healthcare settings and in the Kewot woreda, to understand transmission pathways and inform recommended interventions, such as maternal screening or vaccination for certain organisms. Our study helps to fill a knowledge gap regarding carriage prevalence of key bacterial pathogens among pregnant women and neonates in this region.

## Data Availability

De-identified data will be made publicly available upon publication of the manuscript.

## Acknowledgements

We would like to extend our gratitude to the families who participated in this study and made this work possible. We appreciate the HaSET data collectors and field team for their tireless efforts to successfully implement this study. We are grateful for the support of the HaSET community advisory board. We would also like to thank Anne Redmond Sites and Clara Pons Durans for their helpful comments on the manuscript. Finally, we would like to thank the Whole Genome Sequencing facility at NICD for helping with analysis of data and Ruth Mogokotleng for interpretation.

## Author contributions (based on CRediT Taxonomy)

*Conceptualization:* C.T.W., D.B., G.C.

*Data curation:* G.A., C.T.W., O.P., A.G., M.S.

*Formal analysis:* G.A., C.T.W.

*Funding acquisition:* G.C.

*Investigation:* G.A., C.T.W., O.P., A.G., M.S., G.C.

*Methodology:* G.A., C.T.W., O.P., B.M.H., M.S., D.B., G.C.

*Project administration:* G.A., O.P., A.G., M.S., A.F., G.C.

*Resources:* G.A., O.P., A.G., M.S., A.F., G.C.

*Software:* C.T.W.

*Supervision:* O.P., D.B., L.T., G.C.

*Validation:* G.A., C.T.W., O.P., M.S.

*Visualization:* C.T.W.

*Writing – original draft preparation:* G.A., C.T.W., O.P.

*Writing – review & editing:* All

## Data availability

De-identified data will be made publicly available upon publication of the manuscript.

