## Supplemental material for "Carriage of antimicrobial-resistant Enterobacterales among pregnant women and newborns in Amhara, Ethiopia"

### Supplementary Material

#### Contents

**Supplementary Figure 1.** 2x2 tables comparing carriage of ESBL-producing organisms, CRE, and GBS in mothers during labor/delivery and their neonates.

**Supplementary Table 1.** Primer and probe sequences for *bla_-_*_TEM_*, bla_-_*_SHV_ and *bla_-_*_CTXM_ gene detection.

**Supplementary Table 2.** PCR results among 101 ESBL-producing isolates, as identified by AST. PCR was not conducted for 1 isolate.

**Supplementary Table 3.** Antimicrobial resistance genes detected among 16 CRE-positive isolates.

##### **Supplementary Figure 1.** 2x2 tables comparing carriage of ESBL-producing organisms (a), CRE (b), and GBS (c) in mothers during labor/delivery and their neonates.

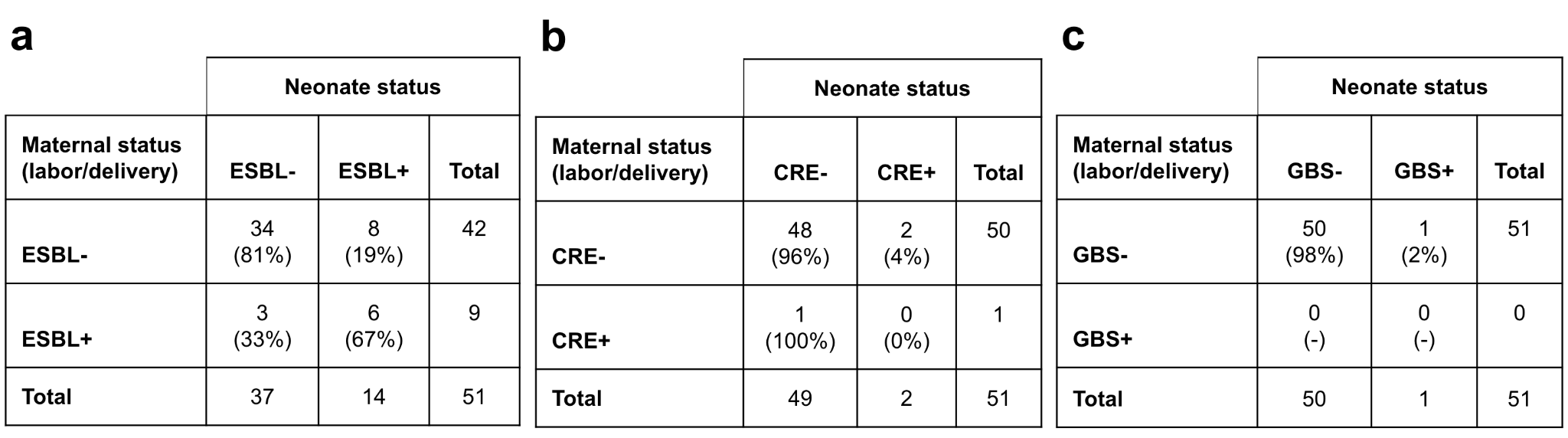

##### **Supplementary Table 1.** Primer and probe sequences for *bla_-_*_TEM_*, bla_-_*_SHV_ and *bla_-_*_CTXM_ gene detection.

| **ESBL primer and probe name** | **Primer and probe sequences** |
| --- | --- |
| TEM Forward Primer | 5’- AAG TTC TGC TAT GTG GTG CGG TA -3’ |
| TEM Reverse Primer | 5’- TGT TAT CAC TCA TGG TTA TGG CAG C -3’ |
| TEM A Primer | 5’- GTA AGA TGC TTT TCT GTG ACT GGT GA -3’ |
| TEM S Primer | 5’- AGT TCT GCT ATG TGG TGC GGT ATT A -3’ |
| TEM Probe | 5’- FAM- TGC GGC GAC CGA GTT GCT CTT –BBQ -3’ |
| SHV Forward Primer | 5’- CAG CAG GAT CTG GTG GAC TAC T -3’ |
| SHV Reverse Primer | 5’- GTC AAG GCG GGT GAC GTT -3’ |
| SHV A Primer | 5’- AAG GCG GGT GAC GTT GTC -3’ |
| SHV S Primer | 5’- CCG GTC AGC GAA AAA CAC -3’ |
| SHV Probe | 5’- Cy5- TCT GGC GCA AAA AGG CAG TCA –BBQ -3’ |
| CTX-M Forward Primer | 5’- ATG TGC AGY ACC AGT AAR GTK ATG GC -3’ |
| CTX-M Reverse Primer | 5’- ATC ACK CGG RTC GCC NGG RAT -3’ |
| CTX-M1 Probe | 5’- FAM- CCC GAC AGC TGG GAG ACG AAA CGT -BBQ -3’ |
| CTX-M2 Probe | 5’- YAK- CAG GTG CTT ATC GCT CTC GCT CTG TT -Q -3’ |
| CTX-M9 Probe | 5’- 610- CTG GAT CGC ACT GAA CCT ACG CTG A –Q - 3’ |
| CTX-M9all/1 Probe (locked nucleic acid [LNA] Probe) | 5’- 640- CG+AC+AAT+ACN GCC+ATG+AA –BBQ -3’ |

##### **Supplementary Table 2.** PCR results among 101 ESBL-producing isolates, as identified by AST. PCR was not conducted for 1 isolate.

| **Gene** | **Count (%)** |
| --- | --- |
| TEM only | 3 (3.0) |
| SHV only | 1 (1.0) |
| CTX-M-1 only | 31 (30.7) |
| CTX-M-9all only | 2 (2.0) |
| TEM + SHV | 1 (1.0) |
| TEM + CTX-M-1 | 43 (42.6) |
| TEM + CTX-M-9all | 4 (4.0) |
| SHV + CTX-M-1 | 6 (5.9) |
| CTX-M1 + CTX-M-9all | 1 (1.0) |
| TEM + SHV + CTX-M-1 | 6 (5.9) |
| None | 3 (3.0) |

##### **Supplementary Table 3.** Antimicrobial resistance genes detected by WGS among 16 CRE-positive isolates.

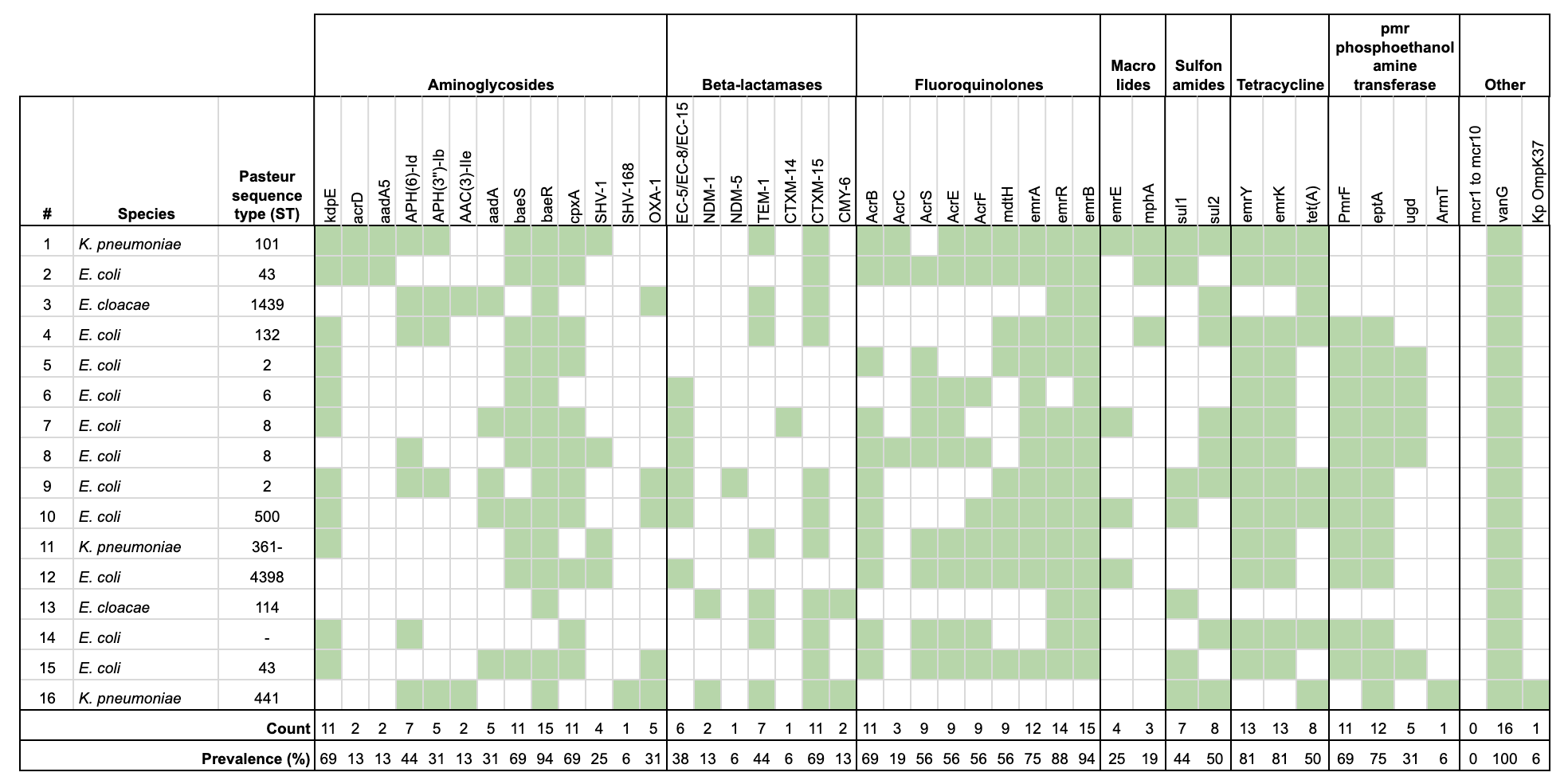
